# The Medication Patterns of Spinocerebellar Ataxia Type 3 Mutation Carriers Enrolled in the ESMI Cohort

**DOI:** 10.1101/2025.03.05.25322961

**Authors:** Patrick Silva, Marina A. Costa, Laetitia Gaspar, João Durães, Inês Cunha, Joana A Ribeiro, Cristina Januário, Bárbara Oliveiros, Jeannette Hübener-Schmid, Jennifer Faber, Mafalda Raposo, Manuela Lima, Hector Garcia-Moreno, Paola Giunti, Lukas Beichert, Ludger Schöls, Bart P. van de Warrenburg, Jeroen de Vries, Andreas Thieme, Kathrin Reetz, Heike Jacobi, Jon Infante, Thomas Klockgether, ESMI Study Group, Luís Pereira de Almeida, Magda M. Santana

## Abstract

**Background:** Spinocerebellar ataxia type 3 (SCA3) is one of the most common dominantly inherited ataxia worldwide. Despite research advances, no approved disease-modifying treatment exists, and management focuses on symptom alleviation and functional capacity maximization. Symptomatic treatment guidelines are scarce, leaving decisions to physicians’ discretion. The lack of studies on SCA3 symptom management hinders therapy standardization. This study investigated medication usage patterns among SCA3 mutation carriers and controls recruited by the multicentric European Spinocerebellar Ataxia Type-3/Machado-Joseph Disease Initiative (ESMI).

**Methods:** We collected medication data from ESMI cohort participants (n=474), comparing SCA3 mutation carriers (n=344) at different disease stages with controls (n=130). We analysed medication usage based on age and ataxia severity groups as well as research centre locations using the ATC code system for classification.

**Results:** There were significant differences in medication usage between mutation carriers and controls. SCA3 subjects took more vitamins, mineral supplements, and muscle relaxants, and medications targeting the nervous system. Psychoanaleptics and vitamins were introduced earlier in the disease course, with 29.2% and 25.0% of mildly ataxic individuals using such subclasses medications, respectively. Most medications, however, were only initiated during the mid-to-late stages of the disease, coinciding with the onset of most neurological symptoms. There were substantial disparities in medication usage across study centres. No significant impact on disease progression was observed for the medication subclasses more frequently used by SCA3 patients.

**Conclusions:** This is the first study to explore medication usage patterns in SCA3 mutation carriers. Our study provides a comprehensive overview of the medications administered in SCA3 and underscore the importance of collaborative efforts toward achieving standardized clinical practices in the management of this disease.

## Introduction

Spinocerebellar ataxia type 3 (SCA3), also known as Machado-Joseph disease (MJD), is an autosomal dominantly-inherited neurodegenerative disease and one of the most prevalent forms of dominant ataxias worldwide (Sullivan et al. 2019; Ruano et al. 2014). The clinical presentation of SCA3 arises from progressive damage to specific neural regions responsible for balance and coordination, particularly the cerebellum, brainstem, and basal ganglia (Rub, Brunt, and Deller 2008; Scherzed et al. 2012). As neurodegeneration advances, patients experience a variety of motor and nonmotor symptoms. Motor symptoms primarily include limb and gait ataxia, with gait impairment and postural coordination deficits often being the initial signs observed in approximately two-thirds of SCA3 cases (Globas et al. 2008). Additionally, individuals can also present oculomotor abnormalities, pyramidal and extrapyramidal features, peripheral nerve involvement, and brainstem dysfunction. Nonmotor symptoms are also prevalent, encompassing sleep disturbances, dysautonomia, cognitive impairment, and vestibular dysfunction (Klockgether, Mariotti, and Paulson 2019; Hengel et al. 2023). These diverse symptoms collectively contribute to the complex clinical presentation of SCA3.

Despite recent advancements in both nonclinical and clinical research, there are currently no approved disease-modifying treatments available for SCA3 (Brooker et al. 2021). Consequently, disease management primarily focuses on symptom alleviation, aiming to maximize patients’ functional capacity and minimize complications associated with the disease progression. Various therapeutic options are employed, such as riluzole and valproic acid to alleviate cerebellar ataxia (yet lacking clinical evidence), antispasmodics to control spasticity, prismatic lenses to correct diplopia, analgesics, psycholeptics and psychoanaleptics to manage pain and mental alterations, among others (Klockgether, Mariotti, and Paulson 2019; de Silva et al. 2019; Zesiewicz et al. 2018; Ilg et al. 2014). However, guidelines for the management of therapeutic interventions in SCA3 or other ataxias are limited, and their utilization is typically based on the discretion of the attending physicians. Currently, no study has systematically reported how symptoms are managed in individuals with SCA3 or any other form of SCA, thereby hindering the standardization of symptom-related therapies and improvement of disease management strategies.

Given the lack of research in this area, we investigated medication patterns for the symptomatic management of SCA3 using a large cohort of mutation carriers from the ESMI cohort. This study provides the first comprehensive insight into the real-world medication patterns in SCA3, aiming to facilitate the development of more standardized and effective treatment approaches to improve the quality of life for patients affected by this debilitating disease.

## Methods

### Ethical approval

This study was conducted by the European Spinocerebellar Ataxia Type 3/Machado-Joseph Disease Initiative (ESMI) following the approval of local ethical committees at all participating centres. In agreement, ethics comittee/IRB of University Hospital of Bonn gave ethical approval for this work. Ethics comittee/IRB of University of Coimbra gave ethical approval for this work. Ethics comittee/IRB of University College London gave ethical approval for this work. Ethics comittee/IRB of Universidade dos Acores, Ponta Delgada gave ethical approval for this work. Ethics comittee/IRB of Radboud University Medical Center gave ethical approval for this work. Ethics comittee/IRB of University Medical Center Groningen gave ethical approval for this work. Ethics comittee/IRB of University of Tuebingen gave ethical approval for this work. Ethics comittee/IRB of University Hospital Essen, University of Duisburg-Essen, gave ethical approval for this work. Ethics comittee/IRB of University Hospital of Heidelberg gave ethical approval for this work. Ethics comittee/IRB of University Hospital Marques de Valdecilla-IDIVAL, Santander, gave ethical approval for this work. Ethics comittee/IRB of RWTH Aachen University gave ethical approval for this work. Informed consent was obtained from all participants prior to enrolment, in accordance with the Declaration of Helsinki.

### Study population and clinical and demographic data collection

The study included individuals with molecularly confirmed SCA3 mutation (*ATXN3* repeat length ≥ 55 CAGs) and control subjects (excluded for the *ATXN3* expansion) from the ESMI cohort. Participants were recruited from 11 research centres: Aachen, Azores, Bonn, Coimbra, Essen, Groningen, Heidelberg, London, Nijmegen, Santander, and Tübingen. Comprehensive demographic, epidemiological, and clinical data were extracted from a shared registry platform.

SCA3 mutation carriers were categorized based on the Scale for the Assessment and Rating of Ataxia (SARA) (Schmitz-Hubsch et al. 2006) score as pre-ataxic (SARA score < 3), mildly ataxic (3.0 ≤ SARA ≤ 7.5), moderately ataxic (8.0 ≤ SARA ≤ 14.5), moderately-severe ataxic (15.0 ≤ SARA ≤ 23.5), or severely ataxic (SARA ≥ 24.0), considering the most recent follow-up study at the date of export (April 21, 2023). The Inventory of Non-Ataxia Signs (INAS) was used to assess non-ataxia manifestations (Jacobi et al. 2013). Stratification by age (three groups: 18-39 years, 40-59 years, and +60 years) and recruitment research centres was also performed.

### Medication data collection

The analysis included data on participants’ ongoing medication usage at their most recent follow-up study visit up to the data export date (performed on April 21, 2023). Participants with missing or incomplete information regarding their current medication status (*i.e.*, whether the medication was ongoing or discontinued) were excluded from the analysis. Medication classified as *as needed* or *seasonal* were excluded from all analyses. Due to considerable missing data on dosage and treatment duration, these variables were not considered for the study. The study followed a cross-sectional design for all the analysis, except to investigate the impact of medication subclasses on disease progression. In this case, a retrospective analysis of longitudinal data was performed.

### Medication classification

All medications were categorized based on the Anatomical Therapeutic Chemical (ATC) classification system up to the second level. This system organizes medication into classes based on the organ or system on which they act. Considering that the same substance (or combination) may receive multiple ATC classifications depending on the prescribed indication, in instances of uncertainty, participants’ comorbidities were assessed to understand the rationale behind the medication usage, and clinicians at each centre were consulted to determine the appropriate ATC code.

A ‘virtual’ ATC subclass, denoted as V90, was established to encompass all unspecified herbal and traditional medicine substances. The following products, despite not having a specified ATC code, were classified as follows: coenzyme Q10 – vitamins (A11), creatine – other alimentary tract and metabolism products (A16), resveratrol – general nutrients (V06), spermidine – other alimentary tract and metabolism products (A16). Genito-urinary system and sex hormones (ATC class G) were excluded from the global analysis due to their sex and age-related use.

### Statistical analysis

Continuous variables (age, age of onset, number of CAG repetitions in mutant allele, disease duration, SARA score, total number of medications) were summarized using medians and interquartile ranges. For inferential statistics involving two populations, the Mann_Whitney *U* test was employed, while the Kruskal_Wallis *H* test was used for comparisons involving more than two groups, considering the nonparametric distribution profiles of all analysed variables. Fisher’s exact test was applied to compare frequencies between groups. A *p*-value less than 0.05 was considered statistically significant. All the statistical analyses were conducted using GraphPad Prism version 9.3.1 (GraphPad Software, San Diego, USA).

To evaluate the association between medication use and disease progression, we fitted linear mixed-effects models (LMMs) using restricted maximum likelihood estimation. The dependent variable was SARA score, modeled as a function of disease duration, medication use (treated vs. non-treated), and their interaction. Models were adjusted for age, SARA score at baseline, and the number of CAG repeats. A random intercept for each individual accounted for within-subject correlation across repeated measures. Participants classified as controls or pre-ataxic individuals (SARA < 3 at their last longitudinal visit) were excluded from the analysis, as their lack of disease progression would not contribute meaningfully to the models. Additionally, three individuals were excluded following a visual inspection of the dataset, as their disease duration and SARA scores were markedly different from the rest of the cohort. Observations with missing data were also excluded to ensure a complete-case analysis. For each medication subclass, we estimated the effect of treatment on disease progression by comparing the slopes of SARA progression over time between treated and non-treated individuals. Pairwise comparisons of disease progression rates (slopes) were conducted using estimated marginal trends. To visually represent disease progression, we generated spaghetti plots showing individual SARA trajectories, overlaid with group-level regression lines derived from the adjusted mixed-effects models. All analyses were conducted in R version 4.4.1 using the “lme4”, “lmerTest”, and “emmeans” packages. Statistical significance was set at p *<* 0.05.

## Results

### A higher percentage of SCA3 mutation carriers are medicated compared to controls, but differences are only found in the younger age strata

A population of 474 participants, consisting of 344 individuals carrying the *ATXN3* mutation (51.2% female; 51.0 median age; 12.0 median SARA score) and 130 control individuals (56.2% female; 45.0 median age; 0.0 median SARA score), was included in this study. Demographics, epidemiology, and clinical information of the study population and stratification groups are summarized in Supplementary Table S1. The largest number of SCA3 participants were recruited in Azores (n=74; 21.5%), followed by Bonn (n=66; 19.2%), Coimbra (n=57; 16.6%), London (n=46; 13.4%), and Tübingen (n=39; 11.3%; Figure 1A). No significant differences in age or disease duration were observed across study centres (Figure 1B). However, mutation carriers from the Azores had significantly longer CAG repeats than those from Bonn and Tübingen, and an earlier age at onset than those from Bonn (median CAG: Azores = 70.0; Bonn = 68.5; Tübingen = 67.0). Additionally, individuals from London exhibited higher SARA scores compared to those from Bonn and Tübingen (median SARA: London = 17.0; Bonn = 8.5; Tübingen = 9.8; Figure 1B, Supplementary Table S2).

**Figure 1.**
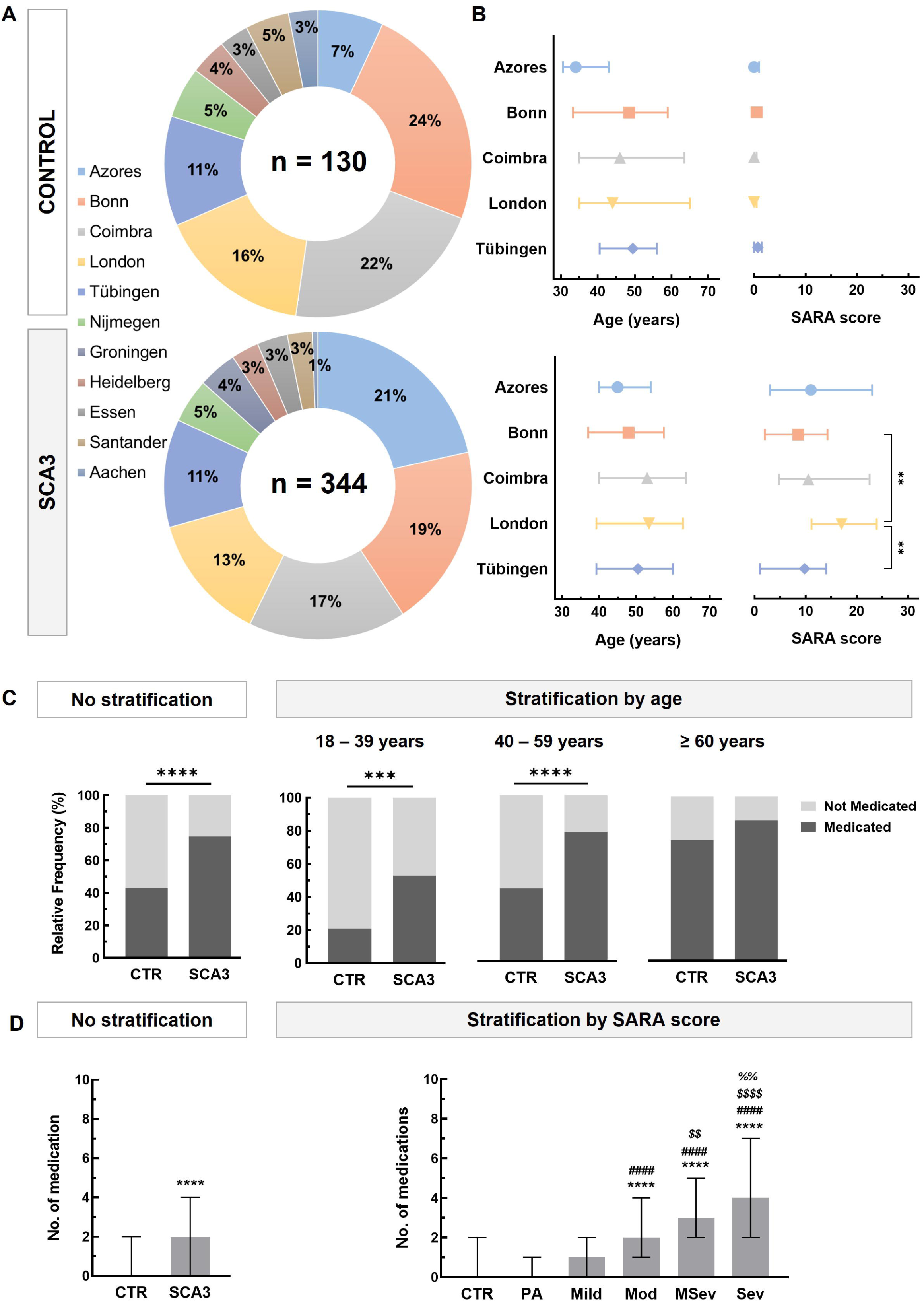
Cohort characterization, frequency of medicated subjects and number of medications per subject within the study population. **A.** Relative distribution of enrolled participants across research centres. Bonn, Azores, Coimbra, London, and Tübingen are the five primary recruiting centres, together accounting for 79% and 82% of control (top) and SCA3 (bottom) study populations, respectively. **B.** Comparative analysis of age (left) and SARA score (right) among control (top) and SCA3 (bottom) study populations from the five primary recruiting centres. SCA3 subjects from London have significantly higher SARA scores than those from Bonn and Tübingen. Symbols represent median values, and error bars indicate the interquartile range. Kruskal_Wallis *H* test, followed by Dunn’s *post-hoc* test for multiple comparisons. \*\**p*<0.01. **C.** Comparative analysis of the relative frequency of medicated subjects in SCA3 and control groups. A higher percentage of SCA3 subjects are medicated compared to control individuals (no stratification). Differences between the two groups are observed for the 18 to 39 years and 40 to 59 years stratification groups, but not in individuals aged 60 or above (stratification by age). Fisher’s exact t-test. \**p*<0.05; \*\*\*\**p*<0.0001. **D.** Number of medications used per subject in control and SCA3 groups, and stratification by SARA score. The severity of ataxia influences the number of medications taken, with subjects in advanced disease stages using significantly more medications than those in the early stages. Data presented as median values and interquartile range. Kruskal_Wallis *H* test, followed by Dunn’s *post-hoc* test for multiple comparisons. \*\**p*<0.01; \*\*\*\**p*<0.0001. * denotes a comparison against controls; # against pre-ataxic individuals; $ against mild ataxic subjects; % against moderately ataxic subjects.

Among SCA3 subjects, 257 individuals (74.7%) were taking at least one medication. In contrast, a significantly lower percentage of controls (43.1%) were medicated (*p*<0.0001). This difference is primarily driven by younger individuals. For participants with 60+ years, the percentage of medicated individuals was comparable in both the control (73.3%) and SCA3 (85.2%) groups. Conversely, in the 40-59 years age group, a greater percentage of SCA3 mutation carriers were medicated (78.0%) than controls (43.9%, *p*<0.0001). Similarly, in the younger age group (18-39 years), more SCA3 mutation carriers were taking medication (52.9%) than controls (20.9%, *p*=0.0008; Figure 1C, Supplementary Table S3). The percentage of participants medicated stratified by SARA score and research centre can be found in Supplementary Table S3.

### SCA3 mutation carriers at more advanced disease stages are significantly more polymedicated

Overall, the number of medications taken by SCA3 subjects (median: 2, IQR [0–4]) was higher than controls (0, [0–2]; *p*<0.0001; Figure 1D). To explore whether the disease stage influences the number of medications taken by SCA3 mutation carriers, we stratified subjects based on their SARA score and compared the number of medications within each group. No statistically significant differences were found between controls and both the pre-ataxic (0, [0–1]) and mild ataxia (1.5, [0–2]) groups, nor between the pre-ataxic and mild ataxia groups. However, individuals with moderate (2, [1–4]) or moderately-severe ataxia (3, [2–5]) were significantly more polymedicated than controls and pre-ataxic individuals (*p*<0.0001 for all comparisons, Figure 1D). Additionally, a significant difference in the number of medications taken was noted between the moderate and moderately-severe ataxia groups (*p*=0.0041). Severe ataxic individuals (4, [2–7]) were significantly more polymedicated than controls, pre-ataxic, mildly ataxic (*p*<0.0001 for all comparisons), and moderately ataxic SCA3 mutation carriers (*p*=0.0094).

### SCA3 mutation carriers take more medication for the nervous system, the alimentary tract and metabolism, and the musculoskeletal system

To identify the medication classes taken by participants, we used the ATC classification system. Medications for the nervous system were the most used among SCA3 subjects, with 159 individuals (46.2%) taking at least one medication from this class. The next most frequently taken medication classes were those for the alimentary tract and metabolism (40.4%), cardiovascular (25.9%), and musculoskeletal (21.5%) systems. Among the control subjects, the most prevalent medication classes were the cardiovascular system (17.7%), the alimentary tract and metabolism (17.7%), and the genito-urinary system and sex hormones (13.8%). Nervous system medications were used by 9.2% of the control group. Comparing the two groups, a significantly higher percentage of SCA3 individuals used medications from the nervous system (*p*<0.0001), alimentary tract and metabolism (*p*<0.0001), and musculoskeletal system (*p*<0.0001) classes (Table 1).

**Table 1.** Frequency of medicated control and SCA3 subjects for each ATC class.

| ATC Classification<br>(1<sup>st</sup> Level) | Control |  | SCA3 |  | Fisher's exact<br>test (*p* value) |
| --- | --- | --- | --- | --- | --- |
|  | n | % med | n | % med |  |
| ***Nervous system [N]*** | **12** | **9.2** | **159** | **46.2** | **<0.0001** |
| 18-39 years old | 1 | 2.3 | 14 | 20.0 | 0.0084 |
| 40-59 years old | 7 | 12.3 | 99 | 53.2 | <0.0001 |
| 60+ years old | 4 | 13.3 | 46 | 52.3 | 0.0002 |
| ***Alimentary tract and metabolism [A]*** | **23** | **17.7** | **139** | **40.4** | **<0.0001** |
| 18-39 years old | 3 | 7.0 | 17 | 24.3 | 0.0224 |
| 40-59 years old | 10 | 17.5 | 85 | 45.7 | <0.0001 |
| 60+ years old | 10 | 33.3 | 37 | 42.0 | 0.5179 |
| Cardiovascular system [C] | 23 | 17.7 | 89 | 25.9 | 0.0692 |
| ***Musculo-skeletal system [M]*** | **7** | **5.4** | **74** | **21.5** | **<0.0001** |
| 18-39 years old | 0 | 0.0 | 9 | 12.9 | 0.0127 |
| 40-59 years old | 2 | 3.5 | 47 | 25.3 | 0.0001 |
| 60+ years old | 5 | 16.7 | 18 | 20.5 | 0.7925 |
| Blood and blood forming organs [B] | 10 | 7.7 | 39 | 11.3 | 0.3106 |
| Genito-urinary system and sex hormones [G] | 18 | 13.8 | 38 | 11.0 | 0.4261 |
| Systemic hormonal preparations [H] | 13 | 10.0 | 22 | 6.4 | 0.2361 |
| Various [V] | 4 | 3.1 | 19 | 5.5 | 0.3428 |
| Respiratory system [R] | 7 | 5.4 | 12 | 3.5 | 0.4301 |
| Sensory organs [S] | 2 | 1.5 | 7 | 2.0 | >0.9999 |
| Antiinfectives for systemic use [J] | 1 | 0.8 | 6 | 1.7 | 0.6796 |
| Antineoplastic and immunomodulating agents [L] | 1 | 0.8 | 6 | 1.7 | 0.6796 |
| Dermatologics [D] | 1 | 0.8 | 3 | 0.9 | >0.9999 |
| Antiparasitic products, insecticides and repellents [P] | 1 | 0.8 | 0 | 0.0 | 0.2743 |

ATC: Anatomical Therapeutic Chemical. Percentages indicate the relative frequency within the specific population, either SCA3 or control. Classes with statistically significant differences are highlighted in bold and italic.

To investigate the impact of age, we conducted a similar comparison within age stratification groups. In younger subjects (18-39 years of age), differences between mutation carriers and controls were observed for the nervous system (20.0% vs. 2.3%, *p*=0.0084), the musculo-skeletal system (12.9% vs. 0.0%, *p*=0.0127), and the alimentary tract and metabolism (24.3% vs. 7.0%, *p*=0.0224) classes. Similar results were observed for individuals aged 40-59 years: a higher percentage of SCA3 subjects were taking medications targeting at the nervous system (53.2% vs. 12.3%, *p*<0.0001), alimentary tract and metabolism (45.7% vs. 17.5%, *p*<0.0001), and musculoskeletal system (25.3% vs. 3.5%, *p*=0.0001) classes, compared to controls. In older participants, differences between SCA3 and controls were found only for the nervous system class (52.3% vs. 13.3%; *p*=0.0002; Table 1). A detailed analysis of each medication class stratified by age, SARA score and research centre is available in Supplementary Table S4.

### SCA3 individuals take more vitamins and mineral supplements, as well as muscle relaxant, psychoanaleptic, anti-Parkinson, analgesic, psycholeptic, and antiepileptic medication

Within the nervous system class, psychoanaleptics are the most frequently used medications, accounting for 25.3% of the SCA3 study population, followed by anti-Parkinson drugs (15.4%), analgesics (14.8%), psycholeptics (13.7%), and antiepileptics (7.0%). Statistically significant differences were observed between the SCA3 study population and controls in the usage of all nervous system subclasses, except for anesthetics and other nervous system drugs subclasses. These include the psychoanaleptics (*p*<0.0001), anti-Parkinson drugs (*p*<0.0001), psycholeptics (*p*<0.0001), analgesics (*p*=0.0002), and antiepileptics (*p*=0.0006; Table 2).

**Table 2.** Frequency of medicated controls and SCA3 subjects for ATC subclasses.

| ATC Classification<br>(2<sup>nd</sup> Level) | Control |  | SCA3 |  | Fisher's exact test<br>(p value) |
| --- | --- | --- | --- | --- | --- |
|  | n | % med | n | % med |  |
| ***Psychoanaleptics [N06]*** | **8** | **6.2** | **87** | **25.3** | **<0.0001** |
| 18-39 years old | 1 | 2.3 | 9 | 12.9 | 0.0861 |
| 40-59 years old | 5 | 8.8 | 56 | 30.1 | 0.0008 |
| 60+ years old | 2 | 6.7 | 22 | 25.0 | 0.0358 |
| ***Vitamins [A11]*** | **7** | **5.4** | **76** | **22.1** | **<0.0001** |
| 18-39 years old | 3 | 7.0 | 14 | 20.0 | 0.1016 |
| 40-59 years old | 2 | 3.5 | 42 | 22.6 | 0.0006 |
| 60+ years old | 2 | 6.7 | 20 | 22.7 | 0.0596 |
| ***Anti-Parkinson drugs [N04]*** | **0** | **0.0** | **53** | **15.4** | **<0.0001** |
| 18-39 years old | 0 | 0.0 | 3 | 4.3 | 0.2865 |
| 40-59 years old | 0 | 0.0 | 30 | 16.1 | 0.0003 |
| 60+ years old | 0 | 0.0 | 20 | 22.7 | 0.0018 |
| ***Analgesics [N02]*** | **4** | **3.1** | **51** | **14.8** | **0.0002** |
| 18-39 years old | 1 | 2.3 | 2 | 2.9 | >0.9999 |
| 40-59 years old | 2 | 3.5 | 36 | 19.4 | 0.0019 |
| 60+ years old | 1 | 3.3 | 13 | 14.8 | 0.1134 |
| ***Psycholeptics [N05]*** | **2** | **1.5** | **47** | **13.7** | **<0.0001** |
| 18-39 years old | 0 | 0.0 | 5 | 7.1 | 0.1546 |
| 40-59 years old | 0 | 0.0 | 33 | 17.7 | <0.0001 |
| 60+ years old | 2 | 6.7 | 9 | 10.2 | 0.7273 |
| ***Mineral supplements [A12]*** | **7** | **5.4** | **46** | **13.4** | **0.0138** |
| 18-39 years old | 1 | 2.3 | 4 | 5.7 | 0.6478 |
| 40-59 years old | 3 | 5.3 | 29 | 15.6 | 0.0453 |
| 60+ years old | 3 | 10.0 | 13 | 14.8 | 0.7583 |
| ***Muscle relaxants [M03]*** | **0** | **0.0** | **41** | **11.9** | **<0.0001** |
| 18-39 years old | 0 | 0.0 | 7 | 10.0 | 0.0429 |
| 40-59 years old | 0 | 0.0 | 29 | 15.6 | 0.0003 |
| Muscle relaxants [M03] | 0 | 0.0 | 41 | 11.9 | <0.0001 |
| ***Antiepileptics [N03]*** | **0** | **0.0** | **24** | **7.0** | **0.0006** |
| 18-39 years old | 0 | 0.0 | 5 | 7.1 | 0.1546 |
| 40-59 years old | 0 | 0.0 | 11 | 5.9 | 0.0718 |
| 60+ years old | 0 | 0.0 | 8 | 9.1 | 0.2000 |

ATC: Anatomical Therapeutic Chemical; % med: percentage of medicated individuals within the specific population, either SCA3 or control. Subclasses are sorted by frequency (highest to lowest) in the SCA3 group.

Regarding the alimentary tract and metabolism class, the differences previously observed between control and SCA3 groups can be attributed to vitamins (20.9% vs. 5.5%; *p*<0.0001), mineral supplements (13.4% vs. 5.4%; *p*=0.0138), and drugs for constipation (3.2% vs. 0.0%; *p*=0.0402) subclasses, although the percentage of SCA3 users for the latter did not exceed 5%. For the musculoskeletal system class, the statistically significant difference is explained exclusively by the intake of muscle relaxants, which was higher in the SCA3 than in the control group (11.9% vs. 0.0%, *p*<0.0001; Table 2).

When stratified by age, statistically significant differences between controls and SCA3 aged 18-39 years were only found for the use of muscle relaxants (*p*<0.0001). Among individuals aged 40-59, significant differences were observed for several medication subclasses: vitamins (*p*=0.0006), mineral supplements (*p*=0.0453), muscle relaxants (*p*=0.0003), analgesics (*p*=0.0019), anti-Parkinson drugs (*p*=0.0003), psycholeptics (*p*<0.0001), and psychoanaleptics (*p*=0.0008). In the 60+ age group, differences between controls and SCA3 were noted only for the anti-Parkinson drugs (*p*=0.0018) and psychoanaleptics (*p*=0.0358) subclasses. A detailed analysis of medication uses among other subclasses, including stratification by age, SARA score and research centre, is presented in Supplementary Table S4.

### Ataxia severity influences medication usage by SCA3 subjects

Upon identification of the predominant ATC classes and subclasses of medication taken by individuals with SCA3, we explored the potential impact of ataxia severity on the medication regimen. In the pre-ataxic stage, except for psychoanaleptics (11.1%), vitamins (7.9%) and mineral supplements (5.7%), medication use from all other subclasses was minimal, each constituting less than 5% of the pre-ataxic individuals (Figure 2A). Upon progression to mild ataxia, a significant increase in the use of psychoanaleptics (30.4%, *p*=0.0146), vitamins (26.1%, *p*=0.0150), and psycholeptics (15.2%, *p*=0.0342) was observed. Among mutation carriers with moderate ataxia, vitamins (20.4%) and psychoanaleptics (17.3%) remained the most commonly used subclasses. Interestingly, when comparing individuals with mild and moderate ataxia, no significant differences were observed in the usage percentage for any of the subclasses. From moderate to moderately-severe ataxia, there was a considerable increase in medication utilization percentages, particularly for muscle relaxants (6.1% vs. 22.7%, *p*=0.0026), analgesics (8.2% vs. 26.7%, *p*=0.0015), psycholeptics (8.2% vs. 20.0*%, p*=0.0401), and psychoanaleptics (17.3% vs. 34.1%, *p*=0.0126). A similar trend was observed when comparing mutation carriers with moderately-severe ataxia to those with severe ataxia; only for mineral supplements and antiepileptics was a higher usage percentage not observed. However, no significant differences were detected between the two groups for any of the subclasses. In the severe ataxia group, psychoanaleptics (37.1%), anti-Parkinson drugs (33.9%), analgesics (32.3%), and vitamins (32.3%) were the most used subclasses. Results for groups comparisons of all medications subclasses are available in Supplementary Table S5.

**Figure 2.**
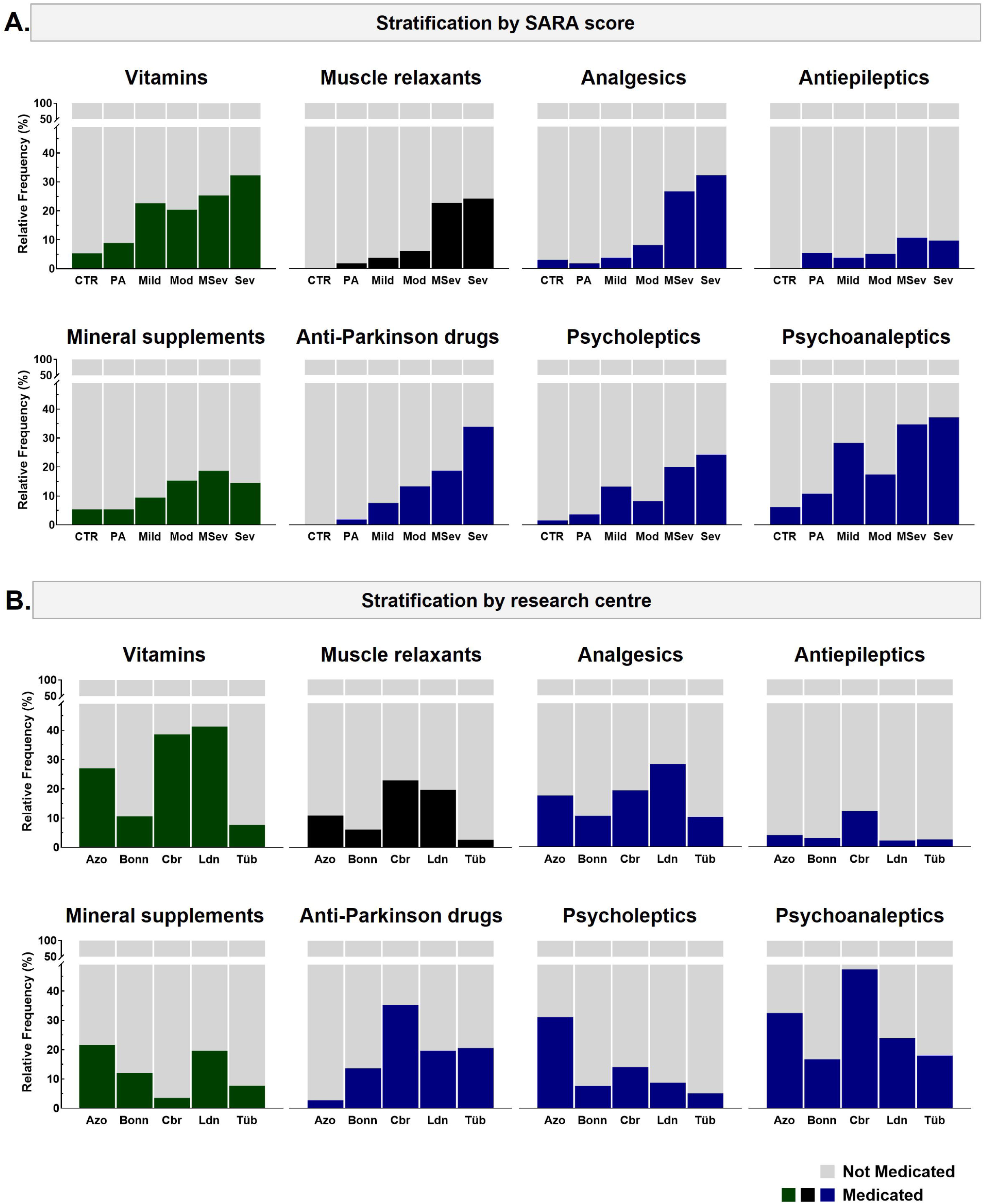
Frequency of medicated subjects for the alimentary tract and metabolism, musculo-skeletal system and nervous system ATC subclasses in SCA3 and control groups, stratified by SARA score and research centre. **A.** Comparative analysis of the relative frequencies of medicated subjects among SCA3 mutation carriers at different stages of ataxia severity, as stratified by SARA score. In the pre-ataxic stage, no significant differences in the relative frequency of medicated subjects were observed compared to the control group for any subclass. The use of vitamins, psycholeptics, and psychoanaleptics markedly increased at phenoconversion. The introduction of analgesics and muscle relaxants is delayed, with significant differences emerging only when comparing moderate-severe ataxic subjects to those with moderate ataxia. Anti-Parkinson drugs use increases gradually with disease progression. Statistical analysis for all these comparisons can be found in Supplementary Table S4. **B.** Comparative analysis of the relative frequencies of medicated subjects among SCA3 mutation carriers recruited and followed at different research centres. The results highlight differences in medication usage patterns across the centres. Mutation carriers from Bonn and Tübingen take significantly less vitamins than those from any other centre, and also less muscle relaxants compared to Coimbra and London subjects. The percentage of subjects using analgesics is lower in London and Bonn cohorts. Coimbra has a high percentage of subjects using antiepileptics and anti-Parkinson drugs, in opposition to the low percentage of mutation carriers from the Azores cohort using these drugs. Conversely, the percentage of subjects from the Azores using psycholeptics is significantly higher compared to any other centre. Coimbra and Azores have a higher percentage of medicated subjects with psychoanaleptics than the other research centres. Statistical analysis for all these comparisons can be found in Supplementary Table S5. Colours in the plots represent ATC classes: green for alimentary and metabolism, black for musculoskeletal system, and blue for nervous system. CTR: control; PA: pre-ataxic; Mild: mild ataxia; Mod: moderate ataxia; MSev: moderate-severe ataxia; Sev: severe ataxia.

### Differences in medication patterns are observed between the research centres

To elucidate the medication patterns among the major recruiting ESMI centres (Azores, Bonn, Coimbra, London, and Tübingen), we analysed the medication subclasses used within each centre’s SCA3 cohort (Figure 2B).

London had the highest percentage of mutation carriers using vitamins (41.3%), which is comparable to Coimbra (38.6%) and Azores (27.0%), with no significant differences noted among these centres. Conversely, vitamin utilization in Bonn and Tübingen was significantly lower, representing 10.6% and 7.7% of the total SCA3 population in these centres, respectively. The use of vitamin supplements varied across centres: Vitamin E was the most frequently used vitamin in Coimbra’s cohort (77%). In opposition, 47% of SCA3 subjects took vitamin D in London, contrasting to 5% of the SCA3 subjects taking this vitamin in Coimbra.

Mineral supplements were most used by participants in the Azores (21.6%) and London (19.6%) cohorts. In contrast, only 3.5% of mutation carriers from Coimbra were using mineral supplements, a significantly lower percentage compared to the Azores and London groups (*p*=0.0038 and *p*=0.0110, respectively). Magnesium was the predominant mineral supplement, accounting for 67% of all mineral supplement users.

Significant differences in the usage of muscle relaxants were observed when comparing Coimbra (22.8%) and London (19.6%) to Bonn (6.1%; *p*=0.0089 vs. Coimbra; *p*=0.0370 vs. London) and Tübingen (2.6%; *p*=0.0066 vs. Coimbra; *p*=0.0184 vs. London). Baclofen was the most used muscle relaxant, representing 63.0% of the individuals taking any medication from this subclass.

No differences in analgesic usage between centres were observed, except for a significantly higher percentage of SCA3 subjects medicated in London (28.3%) compared to Bonn (10.6%; *p*=0.0234). Within the subclass, Pregabalin (43.1% of all analgesics users) and gabapentin (25.5%) were the most utilized medications across all research centres.

The use of antiepileptics among mutation carriers was relatively low, with only 7.0% of the entire cohort taking these medications. There were no significant differences in usage between research centres, although Coimbra had the highest percentage of users (12.3%). Clonazepam was the most used medication, accounting for 58% of all mutation carriers receiving antiepileptics.

Coimbra also had the highest percentage of anti-Parkinson medication usage, with 35.1% of the SCA3 cohort receiving at least one medication from this subclass. In contrast, the Azores cohort had minimal usage, with only 2.7% of mutation carriers using anti-Parkinson medication, significantly lower than Coimbra (*p*<0.0001), and all other centres. Significant differences were also noted between Bonn (13.6%) and Coimbra (*p*=0.0017). London (19.6%) and Tübingen (20.5%) had comparable percentages of patients using anti-Parkinson medication. Amantadine was the most prescribed medication, constituting 32% of all anti-Parkinson medication users, followed by pramipexole (26%).

Azores had the highest percentage of SCA3 subjects using psycholeptics (31.1%), significantly higher compared to Bonn (7.6%; *p*=0.0006), Coimbra (14.0%; *p*=0.0244), London (8.2%; *p*=0.0062), and Tübingen (5.1%; *p*=0.0015). Benzodiazepine derivatives were the most frequently used within this subclass, with alprazolam, lorazepam, and diazepam, accounting for 19%, 15%, and 13% of all psycholeptics used, respectively.

Psychoanaleptics were the most used subclass by mutation carriers in Coimbra (47.4%), Azores (32.4%), and Bonn (16.7%). In Tübingen (17.9%), its usage was only surpassed by anti-Parkinson medication, while in London (23.9%), it ranked third after vitamins and analgesics. Significant differences in usage are observed when comparing Coimbra’s cohort to Bonn (*p*=0.0004), London (*p*=0.232), and Tübingen (*p*=0.0044). Additionally, a significantly larger percentage of mutation carriers used psychoanaleptics in Azores compared to Bonn (*p*=0.0338). Selective serotonin reuptake inhibitors (SSRIs) were the most used medications within this subclass, with sertraline, escitalopram, and citalopram accounting for 21%, 21%, and 16% of all psychoanaleptics users, respectively.

Statistical analysis for all comparisons is available in Supplementary Table S6. The complete list of drugs used in each subclass is displayed in Supplementary Table S7.

### Not all SCA3 subjects who present non-ataxic signs are taking recommended medication

Alongside ataxia, SCA3 mutation carriers present a spectrum of non-ataxic signs and symptoms throughout disease progression. These encompass various neurological manifestations for which therapeutic interventions are available. To explore the use of medications aimed at alleviating these signs and symptoms, we conducted a comprehensive review of established guidelines and relevant literature and identified the recommended medications for each treatable symptom (Supplementary Table S8). We then assessed the percentage of participants with non-ataxic signs taking recommended medications according to our classification.

Urinary dysfunction was the most prevalent manageable non-ataxic manifestations in the SCA3 study population, occurring in 48.5% of individuals. Spasticity (25.7%) and dystonia (16.1%) also had notable prevalence, whereas myoclonus, rigidity, resting tremor, and dyskinesia each occur in less than 10% of the SCA3 cohort (Figure 3A). Among these, rigidity and dystonia were the most frequently treated, with 57.9% and 54.5% of affected individuals receiving recommended pharmacological therapy. Conversely, urinary dysfunction (10.2%), and myoclonus (6.9%) were the least managed symptoms (Figure 3B, Supplementary Table S9).

**Figure 3.**
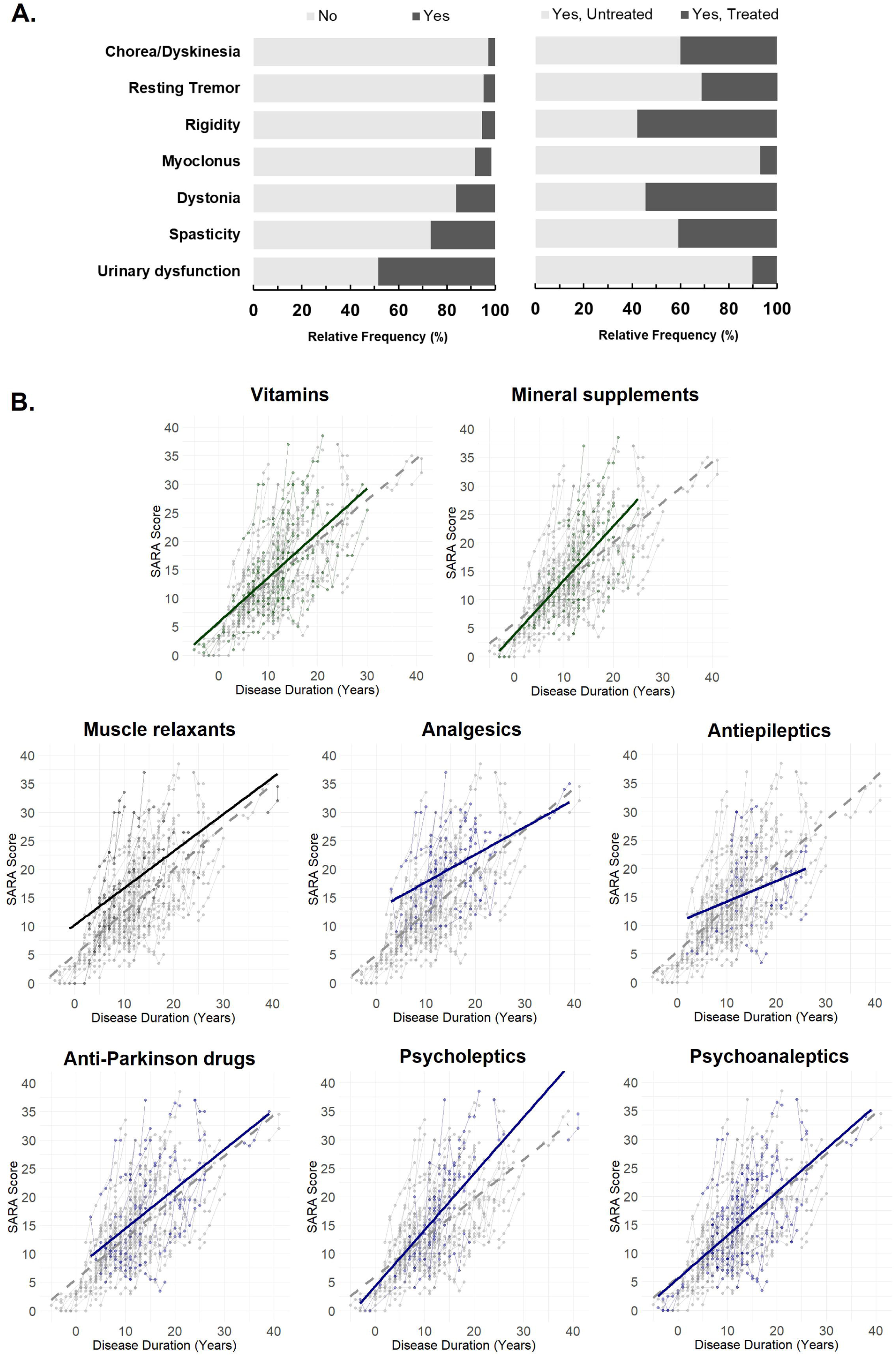
Frequency of manageable non-ataxic signs in SCA3 mutation carriers and impact of medication use on disease progression. **A.** *Left graph.* Stacked bar charts illustrating the relative frequency (%) of SCA3 mutation carriers with (Yes) and without (No) manageable non-ataxic signs, irrespective of age or disease stage. Urinary dysfunction, spasticity, and dystonia are the most frequent signs, while myoclonus, rigidity, resting tremor, and chorea/dyskinesia are less prevalent, with respective frequencies below 10 %. *Right graph.* Stacked bar charts showing the relative frequency (%) of individuals with manageable non-ataxic signs using recommended medication (yes, treated) versus those untreated (yes, untreated). Pharmacological treatment is least frequent for myoclonus and urinary dysfunction, while dystonia and rigidity are the only signs for which more than 50 % of affected individuals receive treatment. **B.** Longitudinal analysis of SARA score in treated and untreated SCA3 mutation carriers across different medication subclasses. Plots depict SARA score trajectories over disease duration (years) for those receiving treatment (solid coloured line) versus untreated individuals (dashed grey line). Mixed-effects linear models were fitted to assess the effect of treatment on the SARA score. No significant differences in SARA score trajectories were observed for vitamins, mineral supplements, muscle relaxants, analgesics, antiepileptics, anti-Parkinson drugs, psycholeptics, or psychoanaleptics.

### None of the medication subclasses more frequently used in SCA3 subjects seem to impact SARA score progression

To assess the effect of medication use on disease progression in SCA3 subjects we took advantage of available retrospective longitudinal data. For each medication subclass we fitted a mixed-effects linear model. In all models, disease duration (β range: 0.302–0.336, all *p*<0.001), baseline SARA score (β range: 0.668–0.694, all *p*<0.001), age (β range: 0.065–0.075, all *p*<0.05), and CAG repeat length (β range: 0.392–0.426, all *p*<0.001) were significant predictors of disease severity (Supplementary Table 10). However, the interaction term between disease duration and treatment status was not significant in any model (*p*>0.05), suggesting that treatment with these medication subclasses does not seem to substantially modify disease progression.

The slopes of SARA score over time were indeed compared between treated and non-treated individuals across different medication subclasses and no meaningful differences in disease progression rates were detected for any subclass: vitamins (Δβ=-0.094, *p*=0.105), mineral supplements (Δβ=-0.122, *p*=0.125), muscle relaxants (Δβ=0.032, *p*=0.627), analgesics (Δβ=0.042, *p*=0.549), antiepileptics (Δβ=0.026, *p*=0.805), anti-Parkinson drugs (Δβ=0.014, *p*=0.823), psycholeptics (Δβ=-0.046, *p*=0.505), and psychoanaleptics (Δβ=0.058, *p*=0.287; Figure 3B, Supplementary Table 11).

## Discussion

At present, there is no curative treatment for SCA3, highlighting the pressing need for effective therapeutic interventions. Current management strategies primarily focus on symptomatic treatments aimed at ameliorating symptoms and enhancing the quality of life for individuals living with ataxia. Symptomatic treatments encompass a range of interventions targeting specific signs and symptoms, such as pain, dystonia, dysautonomia, spasticity, fatigue, rigidity, restless legs syndrome, or oculomotor involvement. Although these treatments do not address the underlying cause of the disease, they play a crucial role in alleviating discomfort and enhancing functional abilities, thereby enhancing the overall well-being and quality of life of individuals affected by the disease. Despite this being true, existing guidelines are scarce, and there is no knowledge of current medication patterns in SCA3, making harmonization between different countries/centres/physicians difficult. This comprehensive analysis of the pharmacological therapies used by SCA3 mutation carriers in the largest European SCA3 cohort has the potential to significantly improve symptomatic management and alleviate the associated burden of the disease by providing clinicians with important information related to real-world medication patterns of SCA3 mutation carriers.

In this study, we showed that a higher percentage of SCA3 mutation carriers are medicated compared to control subjects, as expected. However, this difference is primarily driven by individuals under 60 years old, with the variation in medication usage arising from disease management subclasses, specifically medication targeting the nervous system, vitamins, mineral supplements and muscle relaxants. The increased percentage of medicated elderly controls is likely related to the multimorbidity phenomena associated with aging (Ofori-Asenso et al. 2019), resulting in no significant differences compared to their SCA3 counterpart. Indeed, cardiovascular medication is the most used class among people aged 60 or above in both groups, reflecting typical medical needs associated with aging (Strampelli, Cerreta, and Vucic 2020; Gao et al. 2018).

Our findings suggest that disease management typically begins only when ataxia manifests. Vitamins and psychoanaleptics are often the first subclasses of medication introduced. The general awareness of the benefits of vitamin supplementation (Rai et al. 2021), which usually does not require a prescription, might explain their wide use since early disease, in some way complementing the inexistence of any approved treatment. Of note, although vitamin supplementation is generally perceived as harmless, there are instances where mega-dose intake can lead to neurotoxicity, such as with vitamin B6 (Calderon-Ospina, Nava-Mesa, and Paez-Hurtado 2020), therefore usage should always be advised by a health professional. Reported cases of ataxia with vitamin E deficiency (AVED) (Cavalier et al. 1998; Di Donato, Bianchi, and Federico 2010) might motivate the use of vitamin E, even without any scientific evidence of benefit in SCA3. On its turn, the use of psychoanaleptics since early stages of disease reflect the need to control mood alterations in SCA3 mutation carriers. Reports indicate that depression in patients with neurologic disorders may be attributed to emotional reaction to the diagnosis (Raskind 2008), or are part of the Cerebellar Cognitive Affective Syndrome (CCAS) which may be present in SCA3 mutation carriers (Maas et al. 2021; Thieme et al. 2022; Selvadurai et al. 2024). In SCA3, ataxia progression is associated with a higher risk of depressive syndrome (Schmitz-Hubsch et al. 2011), which is concordant to the continuous high demand of psychoanaleptics in late disease stages we observed. The overall percentage of individuals using psychoanaleptics (25.3%) was consistent with a previous report from the EUROSCA cohort (17.7%, independent of SCA genotype) (Schmitz-Hubsch et al. 2011), and the percentage of SCA3 individuals presenting clinically relevant depression from the CRC-SCA cohort (30.9%) (Lo et al. 2016).

Other subclasses are usually introduced later, with timing seemingly conditioned by various factors, such as the advancement of cerebellar impairment or the emergence of non-ataxic symptoms. Analgesic prescription primarily involves the use of gabapentinoids (gabapentin and pregabalin). Their application in SCAs and other movement disorders is broad, including managing neuropathic pain, spasticity, and restless legs syndrome (de Silva et al. 2019; Benarroch 2021). In SCA3, chronic pain is a frequent and disabling complaint, and is usually a late symptom (Franca et al. 2007), which is consistent with the late introduction of gabapentinoids (and other analgesics) we here report for disease management. Antiepileptics, namely clonazepam, which is classified as an antiepileptic drug, is often employed to manage various neurological symptoms including dystonia, muscle cramps and REM sleep behaviour disorder (de Silva et al. 2019; Termsarasab, Thammongkolchai, and Frucht 2016; Dokkedal-Silva et al. 2020). However, its usage appears to be declining in favour of other subclasses. Interestingly, valproic acid, another antiepileptic previously used in a randomized clinical trial in SCA3 (Lei et al. 2016), is not used in our study population.

Dopaminergic involvement in SCA3 has long been recognized, even at the pre-ataxic stage of the disease (Yen et al. 2000; Yen et al. 2002; Tuite et al. 1995). Parkinsonism has been reported in SCA3, and many other SCAs (Park, Kim, and Jeon 2015), and dopaminergic responses to L-DOPA and dopamine agonists in patients (Bettencourt et al. 2011; Tuite et al. 1995; Buhmann, Bussopulos, and Oechsner 2003) have been supporting the use of dopaminergic agents. Additionally, restless legs syndrome, a frequent symptom among individuals with SCA3 – affecting 17% of ataxic individuals in ESMI cohort (Hengel et al. 2023) –, and a relevant cause of sleep impairment (Schols et al. 1998), is also managed effectively with these anti-Parkinson agents (Scholz et al. 2011; Hornyak et al. 2014).

In general, psycholeptics use is attributed to the management of sleep impairment, which has a substantial impact on the disease burden (Hengel et al. 2023). Benzodiazepines usage can also be helpful to control spasticity, tremor or dystonia (Klockgether, Mariotti, and Paulson 2019; de Silva et al. 2019). On their turn, muscle relaxants, namely baclofen, are recommended for the management of spasticity, dystonia, and rigidity (Termsarasab, Thammongkolchai, and Frucht 2016; Ertzgaard, Campo, and Calabrese 2017), symptoms that often manifest later in the disease course. This justifies the significant increase in its usage among patients at moderate to severe stages of ataxia severity.

We aimed to evaluate whether manageable symptoms were being addressed pharmacologically. Notably, myoclonus and urinary dysfunction were the least treated symptoms, with only 6.9% and 10.2% of affected patients using recommended medication, respectively. For all other evaluated symptoms, except rigidity (57.9%) and dystonia (54.5%), the percentages of mutation carriers using any medication for symptom management were below 50%. The existence of non-pharmacological options for managing many of these symptoms should not be overlooked. For example, physiotherapy is recommended for spasticity (de Silva et al. 2019), while pelvic floor exercises and catheterization can be used for urinary dysfunction. These and other alternatives may influence the actual percentage of individuals receiving treatment when considering non-pharmacological interventions alongside pharmacological treatments. Moreover, we cannot exclude that some medications have been taken by those patients previously without resulting in significant effects and then further discontinued accordingly.

While symptomatic treatments are often prescribed in an attempt to alleviate disease burden, their impact on disease progression remains largely uncertain. In our analysis, no medication subclass demonstrated a significant effect on modifying the rate of SARA progression. This finding suggests that while these treatments may provide symptomatic relief, they do not substantially alter the natural history of the disease. The lack of effect could be attributed to the retrospective nature of the study, potential confounding by indication, or the possibility that these medications simply do not influence disease progression in SCA3.

Besides this longitudinal analysis to assess disease progression, all other analysis result from a cross-sectional design, which inherently provides only a static snapshot of current medication usage at the time of data collection, failing to explore prior medication utilization or reasons for discontinuation. Additionally, all medication information relied solely on participant interviews, which may not accurately reflect prescribed regimens and adherence to treatment protocols. A considerable portion of the dataset lacked crucial details such as initiation dates and dosages, essential for understanding medication indications. Regarding the differences observed between research centres, data interpretation must be performed with caution. Variations in cohort demographics and clinical data between centres might impact the results. Furthermore, participants enrolled in the ESMI cohort are not necessarily clinically managed by the neurologists at the recruitment centre. Information on whether these patients were followed by a neurologist or other physician was lacking but would be interesting to address in future studies. When assessing the usage of recommended medication for the management of specific symptoms, it is important to consider that a given medication may be used to treat multiple overlapping symptoms. This overlap limits the ability to perceive the true extent of symptom-oriented disease management.

In summary, in this study, we investigated the patterns of medication usage among individuals carrying the SCA3 mutation and their control counterparts within the major SCA3-dedicated European cohort. As the first study to explore medication usage profiles in individuals with SCA3, this work offers novel insights into the management of symptomatic SCA3, which might help to develop new or complement existing guidelines. We highlight the following key findings: (1) SCA3 subjects take more vitamins, mineral supplements, muscle relaxants, and medications targeting the nervous system; (2) there are no differences between pre-ataxic and controls in the percentage of participants taking medications for any subclass of medication; (3) many of these medications are only initiated during the mid-to-late stages of the disease, coinciding with the reported onset of many neurological symptoms; (4) there is variability in medication taken by participants across the ESMI study centres; and (5) the medication subclasses that mutation carriers are being medicated with to manage the disease do not seem to have an impact in the progression of ataxia. In summary, our study provides a comprehensive overview of the current medications administered in SCA3. Our findings underscore the importance of collaborative efforts toward achieving standardized clinical practices in the management of SCA3.

## ESMI Study Group

### Ana Ferreira

(Faculdade de Ciências e Tecnologia, Universidade dos Açores, Ponta Delgada, Portugal; UMIB - Unit for Multidisciplinary Research in Biomedicine, ICBAS - School of Medicine and Biomedical Sciences, University of Porto, Porto, Portugal); **Ana Rosa** (Faculdade de Ciências e Tecnologia, Universidade dos Açores, Ponta Delgada, Portugal; UMIB - Unit for Multidisciplinary Research in Biomedicine, ICBAS - School of Medicine and Biomedical Sciences, University of Porto, Porto, Portugal); **Carlos Gonzalez** (Centro de Terapia Familiar e Intervenção Sistémica, Ponta Delgada, Portugal); **Cristina Gonzalez-Robles** (Ataxia Centre, Department of Clinical and Movement Neurosciences, UCL Queen Square Institute of Neurology, London, United Kingdom); **Dagmar Timmann** (Department of Neurology and Hertie-Institute for Clinical Brain Research, University of Tübingen, Tübingen, Germany); **Friedrich Erdlenbruch** (Department of Neurology and Hertie-Institute for Clinical Brain Research, University of Tübingen, Tübingen, Germany); **João Lemos** (Serviço de Psicologia, Hospital do Santo Espírito da Ilha Terceira, Angra do Heroísmo, Portugal); **João Vasconcelos** (Faculdade de Ciências e Tecnologia, Universidade dos Açores, Ponta Delgada, Portugal); **Luís Teves** (Faculdade de Ciências e Tecnologia, Universidade dos Açores, Ponta Delgada, Portugal; UMIB - Unit for Multidisciplinary Research in Biomedicine, ICBAS - School of Medicine and Biomedical Sciences, University of Porto, Porto, Portugal); **Paula Pires** (Serviço de Neurologia, Hospital do Espírito Santo da Ilha Terceira, Angra do Heroísmo, Portugal); **Pedro Lopes** (Serviço de Neurologia, Hospital do Divino Espírito Santo, Ponta Delgada, Portugal); **Pedro Coelho** (Faculdade de Ciências e Tecnologia, Universidade dos Açores, Ponta Delgada, Portugal; UMIB - Unit for Multidisciplinary Research in Biomedicine, ICBAS - School of Medicine and Biomedical Sciences, University of Porto, Porto, Portugal); **Teresa Kay** (Serviço de Genética Clínica, Hospital D. Estefânia, Lisboa, Portugal).

## Data Availability

All data produced in the present study are available upon reasonable request to the authors

## Author’s contribution

Design and conceptualization of the study: PS, MMS, LPA; Subject recruitment/Acquisition of participants data: PS, LG, JD, IC, JAR, CJ, JHS, JF, MR, ML, HGM, PG, LB, LS, BvdW, JdV, AT, KR, HJ, JI, TK, MMS; Statistical analysis of data: PS, MAC, BO, MMS; Drafting of the manuscript: PS, MMS; Revision of the Manuscript: PS, LG, JD, JHS, MR, ML, HGM, LS, BvdW, JdV, AT, KR, HJ, JI, TK, MMS. All authors read and approved the final manuscript.

